# Alcohol Consumption is Associated with Poor Prognosis in Obese Patients with COVID-19: a Mendelian Randomization Study using UK Biobank

**DOI:** 10.1101/2020.11.25.20238915

**Authors:** Xiude Fan, Zhengwen Liu, Kyle L Poulsen, Xiaoqin Wu, Tatsunori Miyata, Srinivasan Dasarathy, Daniel M. Rotroff, Laura E. Nagy

## Abstract

**Background:** Acute and chronic alcohol abuse have adverse impacts on both the innate and adaptive immune response, which may result in reduced resistance to severe acute respiratory syndrome coronavirus-2 (SARS-CoV-2) infection and promote the progression of coronavirus disease 2019 (COVID-19). However, there are no large population-based data evaluating potential causal associations between alcohol consumption and COVID-19.

**Method:** We conducted a Mendelian randomization study using data from UK Biobank to explore the association between alcohol consumption and risk of SARS-CoV-2 infection and serious clinical outcomes in patients with COVID-19. A total of 12,937 participants aged 50-83 who tested for SARS-CoV-2 between 16 March to 27 July 2020 (12.1% tested positive) were included in the analysis. The exposure factor was alcohol consumption. Main outcomes were SARS-CoV-2 positivity and death in COVID-19 patients. We generated weighted and unweighted allele scores using three genetic variants (rs1229984, rs1260326, and rs13107325) and applied the allele scores as the instrumental variables to assess the effect of alcohol consumption on outcomes. Analyses were conducted separately for white participates with and without obesity.

**Results:** Of the 12,937 participants, 4,496 were never or infrequent drinkers and 8,441 were frequent drinkers. (including 1,156 light drinkers, 3,795 moderate drinkers, and 3,490 heavy drinkers). Both logistic regression and Mendelian randomization analyses found no evidence that alcohol consumption was associated with risk of SARS-CoV-2 infection in participants either with (OR=0.963, 95%CI 0.800-1.159; *q* =1.000) or without obesity (OR=0.891, 95%CI 0.755-1.053; *q* =.319). However, frequent drinking (HR=1.565, 95%CI 1.012-2.419; *q* =.079), especially heavy drinking (HR=2.071, 95%CI 1.235-3.472; *q* =.054), was associated with higher risk of death in patients with obesity and COVID-19, but not in patients without obesity. Notably, the risk of death in frequent drinkers with obesity increased slightly with the average amount of alcohol consumed weekly (HR=1.480, 95%CI 1.059-2.069; *q* =.099).

**Conclusions:** Our findings suggested alcohol consumption may had adverse effects on the progression of COVID-19 in white participants with obesity, but was not associate with susceptibility to SARS-CoV-2 infection.

## Introduction

Coronavirus disease 2019 (COVID-19), caused by severe acute respiratory syndrome coronavirus-2 (SARS-CoV-2), is a highly contagious, fast-spreading, and life-threatening infectious disease[1]. So far, it has spread to almost 200 countries and regions, infecting millions of people[2]. In its most serious presentation, COVID-19 can progress rapidly into acute respiratory distress syndrome (ARDS), multi-organ failure, and even death[3, 4]. Thus, identifying potential risk factors for COVID-19 would be a substantial benefit to the public health.

In the midst of the COVID-19 pandemic, off-premise sales of alcohol have increased, according to Nielsen data[5, 6]. Acute and chronic alcohol abuse have adverse impacts on both the innate and the adaptive immune response[7-10] and alcohol consumption is associated with increased susceptibility to pneumonia[9, 11], tuberculosis[9], respiratory syncytial virus (RSV) infection[9, 11], and acute respiratory distress syndrome (ARDS)[9, 12]. Chronic alcohol abuse also exacerbates severity of influenza A virus infection by inhibiting influenza-specific CD8 T cell responses[10]. SARS-COV-2 is a positive-sense, single-stranded RNA (+ssRNA)[13]. Recent data from both murine models of ethanol exposure and peripheral blood mononuclear cells (PBMCs) from patients with alcohol-associated hepatitis (AH) indicate that signaling by viral ss/dsRNA is disrupted by alcohol[14-16], analogous to impact of alcohol on signaling via bacterial products, such as lipopolysaccharide[17]. Therefore, we hypothesized that alcohol consumption may result in reduced resistance to SARS-COV-2 infection and promote the progression of COVID-19.

During the current COVID-19 pandemic, many misconceptions about the protective effects of alcohol in preventing COVID-19 have appeared in social media[18, 19]. Although the World Health Organization (WHO) and other public health authorities have stressed that alcohol consumption does not destroy SARS-CoV-2 and may actually promote infection and accelerate disease progression because of the immunosuppressive effects of alcohol[20], many people around the world still believe that drinking alcohol helps prevent COVID-19[18, 19].

To date, there are no large population-based data evaluating the potential causal associations between alcohol consumption and COVID-19. Therefore, in order to better understand the potential impact of alcohol consumption on the risk of SARS-CoV-2 infection and the progression of COVID-19, we applied the Mendelian randomization approach[21] to evaluate the causal association among participants enrolled in a large national cohort, the UK Biobank, where detailed information on COVID-19, alcohol consumption, other lifestyle factors, comorbidities, and genotype data were rigorously collected.

## Methods

This study is consistent with the Strengthening the Reporting of Observational Studies in Epidemiology (STROBE) guideline. The study did not have a pre-registered or published analysis plan. UK Biobank has obtained Research Tissue Bank (RTB) approval from its ethics committee and this study was also approved by the Institutional Review Boards of the Cleveland Clinic (IRB number:19582).

### Study population from UK Biobank

The UK Biobank, a large population-based prospective cohort, recruited more than 500,000 participants aged 40-69 in 2006-2010 across the United Kingdom. A total of 13,502 participants were tested for COVID-19 between March 16 and July 27, 2020. We excluded participants without alcohol consumption data (n=67) and those without genotype data (n=515). Finally, 12937 participants were included in our study. Participants enrolled in UK Biobank have signed consent forms.

### Exposure of interest

The primary exposure of interest was alcohol consumption. Alcohol consumption data on participants enrolled by the UK Biobank obtained through a self-completed touchscreen questionnaire at the time of enrollment. Participants were asked about their current drinking status (never, previous, current). For current drinkers, they were then asked about the frequency of intake and their average weekly and monthly consumption (the unit is a standard drink) of a range of beverage types (fortified wine, spirits, beer plus cider, red wine, champagne plus white wine).

According to the NIAAA’s classification criteria[22], we classified participants as heavy drinkers (>7 drinks per week for women; >14 drinks per week for men), moderate drinkers (4-7 drinks per week for women; 4-14 drinks per week for men), light drinkers (3 drinks or fewer per week), and never or infrequent drinkers (special occasions only or 1-3 times a month). We also classified heavy drinkers, moderate drinkers and light drinkers as frequent drinkers and those who never drank or drank infrequently as non-drinkers.

### Genetic data

UK Biobank released genetic sequence data from 488,377 individuals genotyped for 847,441 genetic variants in July 2017. Participants were genotyped on either the UK BiLEVE genotyping array (n = 49,950; 807,411 markers) or the UKB Axiom Array (n = 438,427; 825,927 markers). After filtering for genetic variants available on both genotyping arrays and sample quality control processes [23], 488,377 participants with 805,426 single nucleotide variants were available in the release. Detailed information about genotyping and imputation in the UK Biobank has been described previously[23].

Previous genome-wide association study (GWAS) of alcohol consumption in white British participants of UK Biobank identified 14 single nucleotide polymorphisms (SNPs) associated with alcohol consumption[24]. In addition, a study using data from UK Biobank and Genetic Epidemiology Research in Adult Health and Aging (GERA) datasets identified 6 SNPs (Alcohol Dehydrogenase 1B (ADH1B, rs1229984); Klotho Beta (KLB, rs13130794); Basic Transcription Factor 3 Pseudogene 13 (BTF3P13, rs144198753); Glucokinase Regulator (GCKR, rs1260326); Solute Carrier Family 39 Member 8 (SLC39A8, rs13107325); Dopamine Receptor D2 (DRD2, rs11214609)) significantly associated with alcohol consumption[25].

The variant rs1229984 has been successfully used as an instrumental variable in the causal estimation of alcohol consumption and assortative mating[26] and chronic widespread pain in the participants of UK biobank[27]. However, the use of this single SNP may not adequately explain genetic variance in alcohol consumption because of the low minor allele frequency (2.2%) of rs1229984 in the UK Biobank[26]. In order to overcome the potential for weak instrument bias, we generated an allele score utilizing three SNPs: rs1229984 (ADH1B), rs1260326 (GCKR), and rs13107325 (SLC39A8), based on the results of the previous GWAS[25]. These three SNPs were directly genotyped on both the UK BiLEVE and UKB Axiom Arrays and the missingness of these SNPs in participants selected in this study less than 1%. Three additional SNPs (rs13130794 (KLB), rs144198753 (BTF3P13), and rs11214609 (DRD2) were identified in a previous GWAS[25] and were excluded here because they were not directly genotyped (805,426 markers) on the UK Biobank arrays. Detailed information about the three selected SNPs included in this study is shown in **Table S1**. The allele score was calculated per individual as the weighted or unweighted sum of the number of fast alcohol metabolizing alleles of each SNP, whereas the effect of each SNP on alcohol consumption provided in the GERA database (**Table S1**) was used as weight for the calculation of weighted allele score [25, 28].

### Other potential confounding risk factors for COVID-19

A number of potential risk factors for COVID-19 were obtained through the touchscreen questionnaire, inpatient hospital, death register, and genotype data: age at time of COVID-19 test, sex, race (classed as white and non-white ethnic background), body mass index (BMI), blood type, smoking status (no, only occasionally, most or all days), comorbidities (alcohol related diseases, upper gastrointestinal diseases, chronic lower respiratory diseases, chronic heart diseases, diabetes mellitus, dementia, liver cirrhosis and/or liver failure, renal failure, tumor, and acquired immunodeficiency syndrome (AIDS)). In addition, BMI was categorized into four groups according to the WHO classification[29]: underweight (<18.5 kg/m^2^), normal weight (18.5–24.9 kg/m^2^), overweight (25–29.9 kg/m^2^), and obesity (≥30 kg/m^2^).

ICD-10 codes (International Classification of Diseases, Tenth Revision) were used to identify comorbidities and the cause of death from medical records and death records. Alcohol use disorder, alcohol liver diseases, alcohol pancreatitis, alcoholic gastritis, alcoholic cardiomyopathy, alcoholic psychosis, alcoholic myopathy, alcoholic polyneuropathy, and degeneration of the nervous system due to alcohol were uniformly classified as alcohol related diseases. Upper gastrointestinal disease events were defined as participants with gastroesophageal reflux disease (GERD), esophagitis, gastritis/duodenitis, or peptic ulcer. Chronic obstructive pulmonary disease (COPD), asthma, emphysema, and bronchitis/bronchiectasis were uniformly classified as chronic lower respiratory diseases events. Chronic cardiac events were defined as participants with hypertensive, chronic ischemic heart disease, or heart failure. All disease-related ICD 10 codes are shown in the **Table S2**.

### Ascertainment of outcomes

The primary outcome was rate of positive SARS-CoV-2 tests, the secondary outcome was the mortality in COVID-19 positive patients. Data on SARS-CoV-2 test results provided by the UK Biobank covered England, but not Scotland and Wales. Follow-up for mortality was conducted to June 27, 2020 through linkage from National Death Registries. In this study, we selected participants from England in the database and defined the occurrence of outcomes as 0 = non-occurrence, 1= occurrence. COVID-19 death event (n=287) was collected through latest death record with ICD10 code U071 and other deceased COVID-19 positive patients without the corresponding code.

### Statistical analysis

All analyses were performed using Stata (Version 14.0; Stata Corp, College Station, TX). Weekly alcohol consumption was natural log-transformed to meet parametric assumptions. Categorical variables were tested for association using Chi-squared test or Fisher’s exact test if more than 20% of cells had expected frequencies < 5. For multiple comparisons correction, the false discovery rate (FDR) was calculated using the Benjamini-Hochberg method[30] and an adjusted P value (*q-value*) < 0.1 and an unadjusted P value < 0.05 were considered significant.

Alcohol consumption was classified in three ways: (1) A four-level exposure categorical variable including never or infrequent drinkers, light drinkers, moderate drinkers, and heavy drinkers; (2) a binary exposure variable comparing non-drinkers to frequent drinkers; and (3) a log-transformed continuous variable of weekly self-reported alcohol consumption among frequent drinkers.

To reduce the potential confounding effects of factors other than alcohol intake on outcomes, propensity score matching (PSM)[31] was applied to match non-drinkers and frequent drinkers without replacement when we evaluated the associations between alcohol consumption and outcomes. We included variables[32] previously reported to be associated with a higher risk of COVID-19 as the matching factors for PSM. Factors including age, sex, BMI categories, current smoking status, alcohol related diseases, asthma, emphysema, COPD, bronchitis/bronchiectasis, esophagitis, gastritis/duodenitis, peptic ulcer, GERD, hypertensive, chronic ischemic heart disease, heart failure, diabetes, dementia, renal failure, liver cirrhosis and/or liver failure, tumor and AIDS.

We applied two approaches to evaluate the relationship of alcohol consumption with outcomes: logistic or Cox regression association analysis and Mendelian randomization analysis[21]. The logistic regression was applied to evaluate the relationship between alcohol intake and the odds of SARS-CoV-2 infection. The Cox regression analysis was applied to evaluate the relationship between alcohol consumption and the risk of death in COVID-19 positive patients.

We used Mendelian randomization based on three assumptions: (1) the instrumental variable (weighted or unweighted allele score) would not be associated with potentially confounding factors of the risk of COVID-19; (2) the instrumental variable was significantly associated with exposure factor-alcohol consumption; (3) the instrumental variable was only associated with the outcomes through the exposure of interest. We evaluated these three assumptions using linear or logistic regression.

For the Mendelian randomization analysis, the two-stage residual inclusion (2SRI) method[21] was used to calculate the causal effect of alcohol consumption on the outcomes, using the weighted or unweighted allele score as the instrumental variables. The 2SRI method requires adjustment due to different classification methods and variable types (binary or continuous variable) of alcohol consumption and outcomes. For the association of alcohol intake with the risk of SARS-CoV-2 infection, the adjusted 2SRI method was conducted. In the first stage, alcohol intake was associated with the instrumental variable using a logistic regression model (binary exposure of non-drinkers and frequent drinkers) or a linear regression (continuous variable of weekly alcohol consumption in frequent drinkers). In the second stage, the outcome was fit using residuals from the first stage in a logistic regression model. For the risk of death in COVID-19 positive patients, the residuals from the first stage were included as covariates in a Cox regression model. Since the 2SRI method was not suitable for evaluating the association between the four-level categorical variable of alcohol consumption and outcomes, we only applied the logistic or Cox regression analysis for these associations. In addition, we found obesity and race were associated with both the selected SNPs and the risk of COVID-19 (all *q-value*<.10). These analyses were conducted separately for participants with and without obesity, and in those of self-reported white ethnicity.

### Sensitivity analysis

We performed two sensitivity analyses to evaluate the impact of specific subgroup of participants and different definitions of outcomes, respectively, on our analysis. First, in order to clarify the relationship between alcohol intake and outcomes in overweight, but not obese, patients, we selected overweight participants for the sensitivity analysis. Second, because some COVID-19 positive patients were admitted to the intensive care unit (ICU) before they died or recovered and there was no data on life support for patients other than those in ICU, we considered ICU admission and death as serious clinical events and applied logistic regression and Mendelian randomization analyses to evaluate the association between alcohol intake and the risk of severe clinical outcomes.

## Result

### Characteristics of participants

A total of 12,937 participants who had been tested for COVID-19 (from March 16 to July 27, 2020) were selected for our analysis according to the inclusion and exclusion criteria. Of these participants, 1,570 (12.1%) tested positive for COVID-19 and 11,367 (87.9%) tested negative. Regarding the drinking status of these participants, 4,486 (34.8%) were never or infrequently drinkers, 1156 (8.9%) were light drinkers, 3,795 (29.3%) were moderate drinkers, and 3,490 (27.0%) were heavy drinkers. As shown in **Table S3**, significant differences were observed in age, sex, race, blood type, current smoking, and comorbidities other than dementia, tumor, and AIDS between the non-drinkers and frequent drinkers (all *q* <.10). Frequent drinkers tended to have a higher proportion of participants who were older than 65 years (72.0% vs. 69.3%), white (96.6% vs. 86.1%), and of normal weight (28.6 % vs. 24.2%) compared to non-drinkers. In contrast to the known deleterious health effects of heavy drinking [7-10], frequent drinkers had a lower rate of comorbidities than non-drinkers (all *q* <.10). Since the participants in the UK Biobank are between the ages of 50 and 83, we hypothesized that there may be survivor bias between exposure factor (alcohol consumption) and multiple complications. Therefore, to reduce the potential for survivor bias and the effects of confounding factors on outcomes, all analyses were performed in the PSM cohorts and the 2SRI method applied to calculate the causal effect of alcohol consumption on the outcomes [33].

### Instrumental variable associations

Characteristics of participants by rs1229984, rs1260326, and rs13107325 genotypes are shown in **Table S4**. Since the number of participants with alleles rs1229984 and rs13107325, associated with rapid ethanol metabolism, was relatively small, we combined participants with one or two rapid metabolism alleles for comparison. Participants with one or two rapid metabolism alleles compared with those with two reference alleles tended to have a higher proportion of white participants and patients with obesity (all *q* <.10). No significant differences in age, sex, blood types, current smoking, or comorbidities were observed (all *q* <.10).

White Participants with one or two rapid metabolism alleles were less likely to be heavy-drinkers and on average drank less alcohol if they were frequent drinkers (**Table 1 and Figure S1**). These associations are consistent, but the difference between those with 2 or 3 more alleles is greater than those between 0 and 1 (Figure 1). Associations of rs1229984, rs1260326, rs13107325 genotypes, and the weighted or unweighted allele score with alcohol consumption were similar and significant (all *P* <.05). The weighted allele score (F-test=26.289) was more strongly related to alcohol consumption compared with other single SNPs and the unweighted allele score. Furthermore, we did not find an association between SNPs or allele scores and the odds of SARS-CoV-2 infection, the risk of severe clinical outcomes or death in patients with COVID-19 (**Table S5**).

**Table 1.** Association of genetic variations of ADH1B/SLC39A8/GCKR with alcohol consumption in white participants.

| Genetic variations | Alcohol consumption (standard drink/weekly) in frequent drinkers (95% CI) (n=8131) | OR of being a heavy drinker in the whole cohort (95% CI) (n=11982) |
| --- | --- | --- |
| **ADH1B one or two fast alleles vs. none** | -2.777(-4.081- -1.473) | 0.525(0.425-0.647) |
| F-test | 17.424 | — |
| P-value | <0.001 | <0.001 |
| **SLC39A8 one or two fast alleles vs. none** | -0.919(-1.711- -0.127) | 0.874(0.778-0.981) |
| F-test | 5.168 | — |
| P-value | <0.001 | 0.022 |
| **GCKR one or two fast alleles vs. none** | -0.789(-1.360- -0.218) | 0.917(0.845-0.996) |
| F-test | 7.333 | — |
| P-value | <0.001 | 0.039 |
| **Unweighted allele score** | -0.726(-1.066- -0.385) | 0.891(0.848-0.936) |
| F-test | 17.465 | — |
| P-value | <0.001 | <0.001 |
| **Weighted allele score** | -15.068(-20.828- -9.307) | 0.048(0.020-0.118) |
| F-test | 26.289 | — |
| P-value | <0.001 | <0.001 |

**F-test** was calculated by linear regression analysis.
**Abbreviation:** OR, odds ratio; CI, confidence interval; BMI, body mass index.

**Figure 1.**
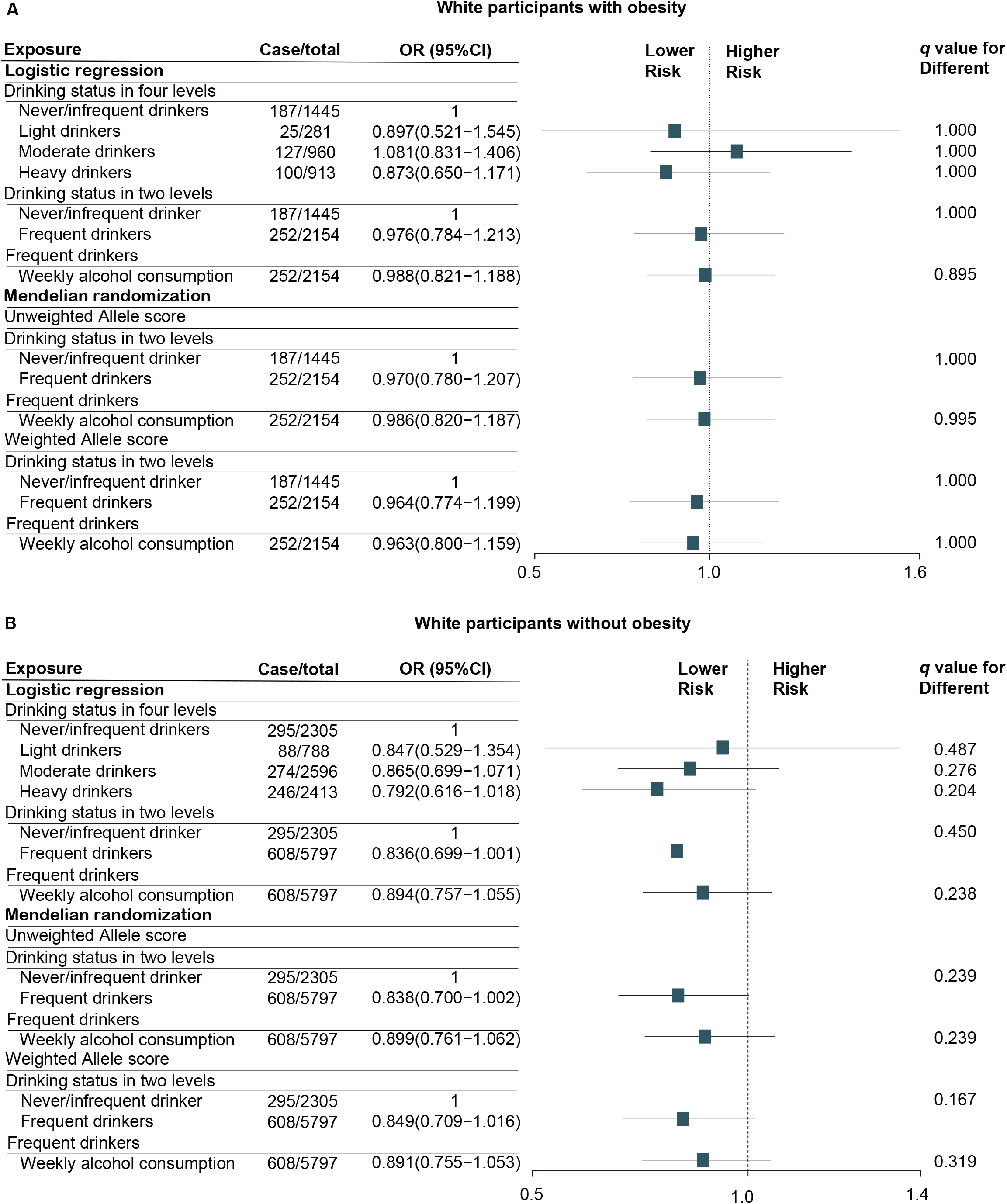
Logistic regression and Mendelian randomization analyses of the association of alcohol consumption with the risk of SARS-CoV-2 infection in white participants with (A) and without obesity (B). Analyses were performed in PSM cohort. Matching factors for PSM including age, sex, BMI categories, current smoking status, alcohol related diseases, asthma, emphysema, COPD, bronchitis/bronchiectasis, esophagitis, gastritis/duodenitis, peptic ulcer, GERD, hypertensive, chronic ischemic heart disease, heart failure, diabetes, dementia, renal failure, liver cirrhosis and/or liver failure, tumor and AIDS. q-value was calculated by false discovery rate (FDR) method. Abbreviation: OR, odds ratio; HR, hazard ratio; CI, confidence interval; PSM, propensity score matching; BMI, body mass index; GERD, gastroesophageal reflux disease; COPD, chronic obstructive pulmonary disease; AIDS, acquired immunodeficiency syndrome.

### Observational associations and instrumental variable associations

The SARS-CoV-2 test positivity rate in white participants was 11.4% (1,368/11,982) and the mortality rate of white patients with COVID-19 was 18.9% (258/1,368). After 1:1 PSM, 1,445 non-drinkers and matched frequent drinkers from the obese cohort and 2,305 non-drinkers and matched frequent drinkers from the non-obese cohort were selected for evaluating the associations between alcohol consumption and the odds of SARS-CoV-2 infection. In addition, 187 non-drinkers and matched frequent drinkers from the obese cohort and 295 non-drinkers and matched frequent drinkers from the non-obese cohort were selected for evaluating the association between alcohol consumption and the risk of worse clinical outcomes of COVID-19. The association of alcohol consumption with the odds of SARS-CoV-2 infection (**Figure 1**) and the risk of death in COVID-19 positive patients (**Figure 2)** are shown.

**Figure 2.**
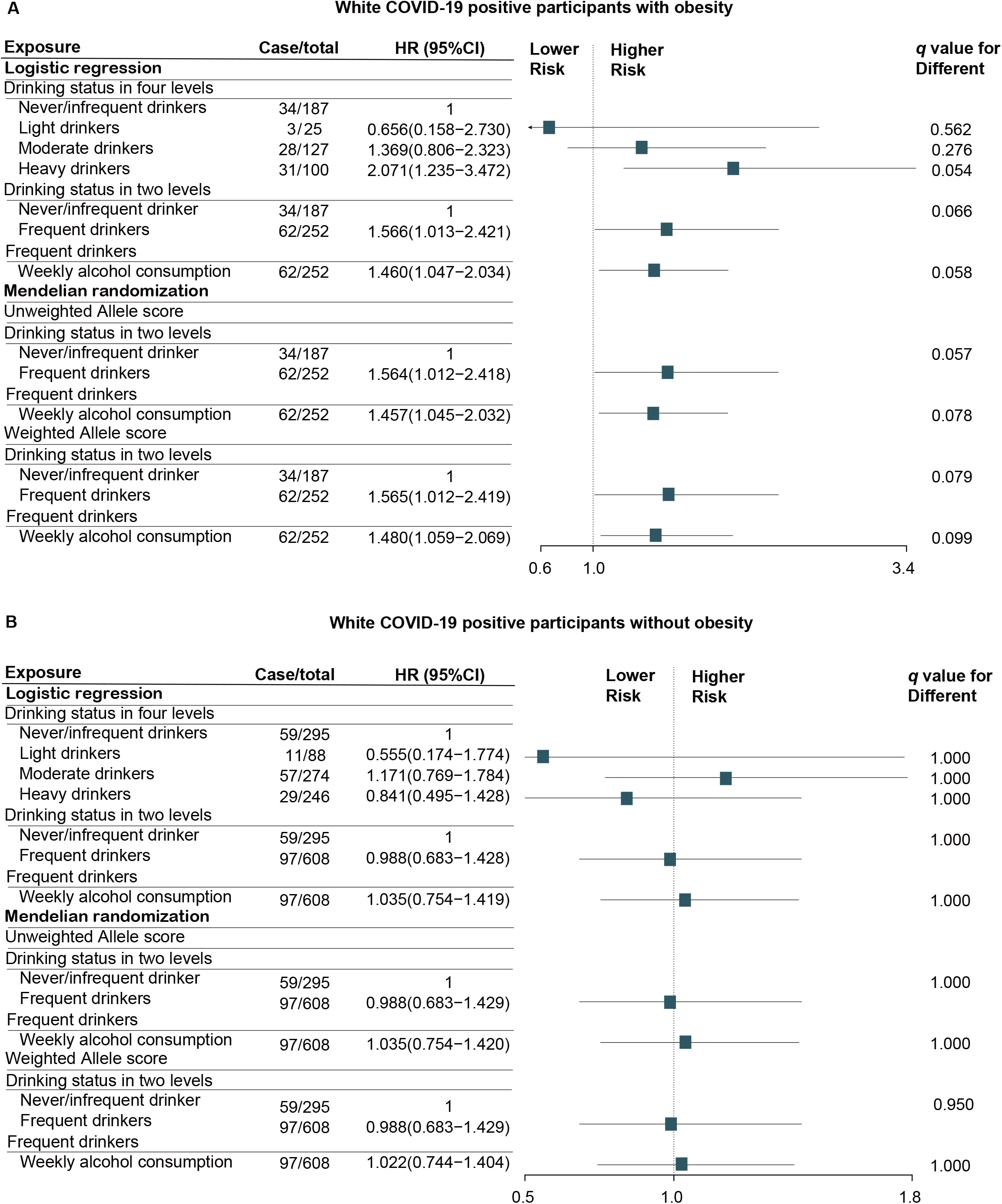
Cox regression and Mendelian randomization analyses of the association of alcohol consumption with the risk of death in white COVID-19 positive patients with (A) and without obesity (B). Analyses were performed in PSM cohort. Matching factors for PSM including age, sex, BMI categories, current smoking status, alcohol related diseases, asthma, emphysema, COPD, bronchitis/bronchiectasis, esophagitis, gastritis/duodenitis, peptic ulcer, GERD, hypertensive, chronic ischemic heart disease, heart failure, diabetes, dementia, renal failure, liver cirrhosis and/or liver failure, tumor and AIDS. q-value was calculated by false discovery rate (FDR) method. Abbreviation: OR, odds ratio; HR, hazard ratio; CI, confidence interval; PSM, propensity score matching; BMI, body mass index; GERD, gastroesophageal reflux disease; COPD, chronic obstructive pulmonary disease; AIDS, acquired immunodeficiency syndrome.

### Alcohol and risk of SARS-CoV-2 infection

For the primary outcome, both logistic regression and Mendelian randomization analyses suggested that alcohol consumption within all three classifications of drinkers --the four-level categorical variable, the binary variable (non-drinkers and frequent drinkers), and the continuous variable of weekly alcohol intake in frequent drinkers was not associated with the risk of SARS-CoV-2 infection (All *q* >.10, **Figure 1**). No association was detected in white participants either with or without obesity. In addition, there was no association between average weekly alcohol consumption and the risk of SARS-CoV-2 infection in either obese (OR=1.065, 95%CI 0.916-1.238; *P*=.412) and non-obese (OR=0.955, 95%CI 0.864-1.057; *P*=.375) cohort before PSM from Mendelian randomization analysis using the weighted allele score.

### Alcohol and risk of death in COVID-19 positive patients

COVID-19 positive patients who were heavy drinkers with obesity had a higher risk of death (HR=2.071, 95%CI 1.235-3.472; *q* =.054, **Figure 2A**). Both Cox regression (HR=1.566, 95%CI 1.013-2.421; *q* =.066) and Mendelian randomization analyses using unweighted allele score (HR=1.564, 95%CI 1.012-2.418; *q* =.057) or weighted allele score (HR=1.565, 95%CI 1.012-2.419; *q* =.079) identified that COVID-19 positive patients with obesity who reported consuming alcohol weekly were more likely to die compared with those drinking none or infrequently. In addition, we found higher alcohol consumption in frequent drinkers resulted in higher risk of death when analyzed by either Cox regression (HR=1.460, 95%CI 1.047-2.034; *q* =.058) or Mendelian randomization analyses using the unweighted allele score (HR=1.457, 95%CI 1.045-2.032; =.078) or the weighted allele score (HR=1.480, 95%CI 1.059-2.069; *q* =.099). However, these associations did not exist in non-obese patients with COVID-19 (*q* >.10, **Figure 2B**). Consistent results regarding the relationship between average weekly alcohol intake and the risk of death was found in both obese (HR=1.418, 95%CI 1.051-1.914; *P*=.022) and non-obese (HR=0.952, 95%CI 0.755-1.201; *P*=.680) cohorts before PSM from Mendelian randomization analysis using weighted allele score. As shown in **Figure 3**, Kaplan–Meier survival plots illustrated that heavy drinkers with obesity had a higher mortality than non-drinkers (Log rank P value=.027), which was not observed in non-obese patients with COVID-19 (Log rank P value=.471).

**Figure 3.**
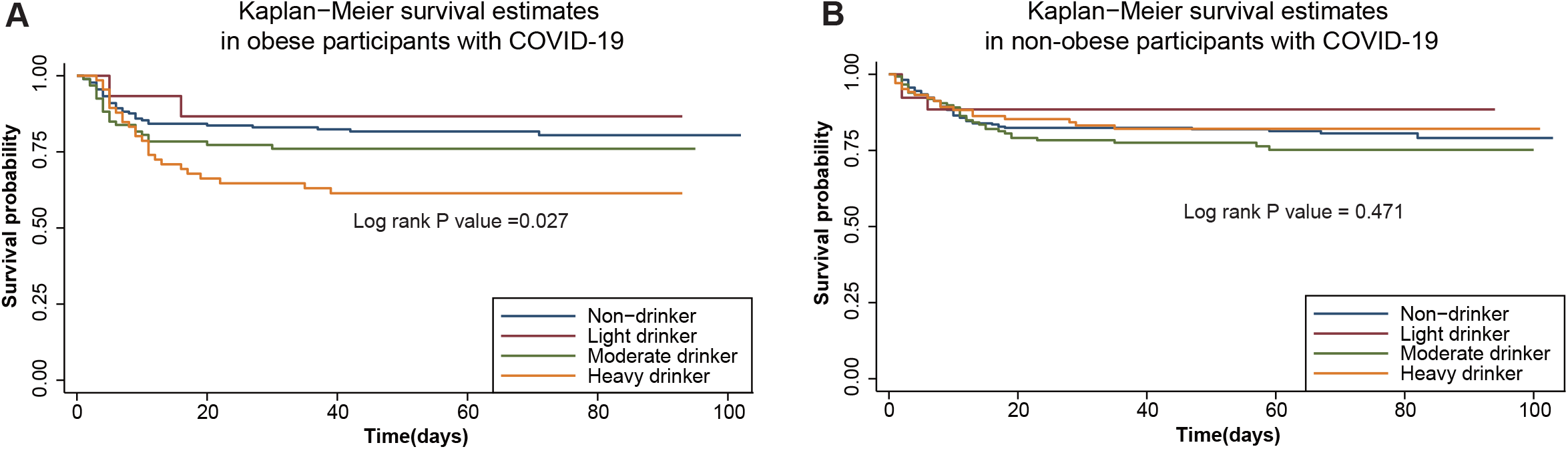
Survival probability of COVID-19 positive patient based on different drinking status in obese (A) and non-obese (B) groups.

### Sensitivity analysis

#### Association between alcohol consumption and outcomes in overweight but not obese patients

A total of 4,869 overweight white participants were tested for SARS-CoV-2. The test positivity rate was 11.7% and the mortality rate of overweight patients with COVID-19 was 18.1% (103/568). Consistent with previous results for patients without obesity (**Figure 1-2**), alcohol consumption was not associated with the risk of SARS-CoV-2 infection or death in overweight COVID-19 positive patients (*q* >.10, **Table S6**).

#### Risk of severe clinical outcomes in COVID-19 positive patients

ICU admission and death in COVID-19 positive patients were grouped together as having severe clinical outcomes. Because some of the patients were admitted to ICU before they were diagnosed with the SARS-CoV-2 infection, we couldn’t use the Cox regression model to analyze the association between alcohol consumption and severe clinical outcomes in the COVID-19 positive cohort without time-to-event data. Heavy drinkers with obesity had a higher likelihood of admission to ICU and death compared to non-drinkers (OR=2.432, 95%CI 1.345-4.397; *q* =.027, **Figure 4A**). Both logistic regression (OR=1.766, 95%CI 1.114-2.801; *q* =.072) and Mendelian randomization analyses using unweighted allele score (OR=1.762, 95%CI 1.11-2.794; *q* =.048) or weighted allele score (OR=1.710, 95%CI 1.077-2.715; *q* =.052) identified that COVID-19 positive patients with obesity who reported consuming alcohol weekly were more likely to suffer severe clinical outcomes compared with those drinking none or infrequently. In addition, we found that the likelihood of serious clinical outcomes in frequent drinkers with obesity slightly increased with the average amount of alcohol consumed weekly based on the result of Mendelian randomization analysis using unweighted allele score (OR=1.020, 95%CI 1.003-1.038, *q* =.095) and weighted allele score (OR=1.018, 95%CI 1.001-1.036; *q* =.054, **Figure 4A**). However, these associations did not exist in non-obese patients with COVID-19 (*q* >.10, **Figure 4B**).

**Figure 4.**
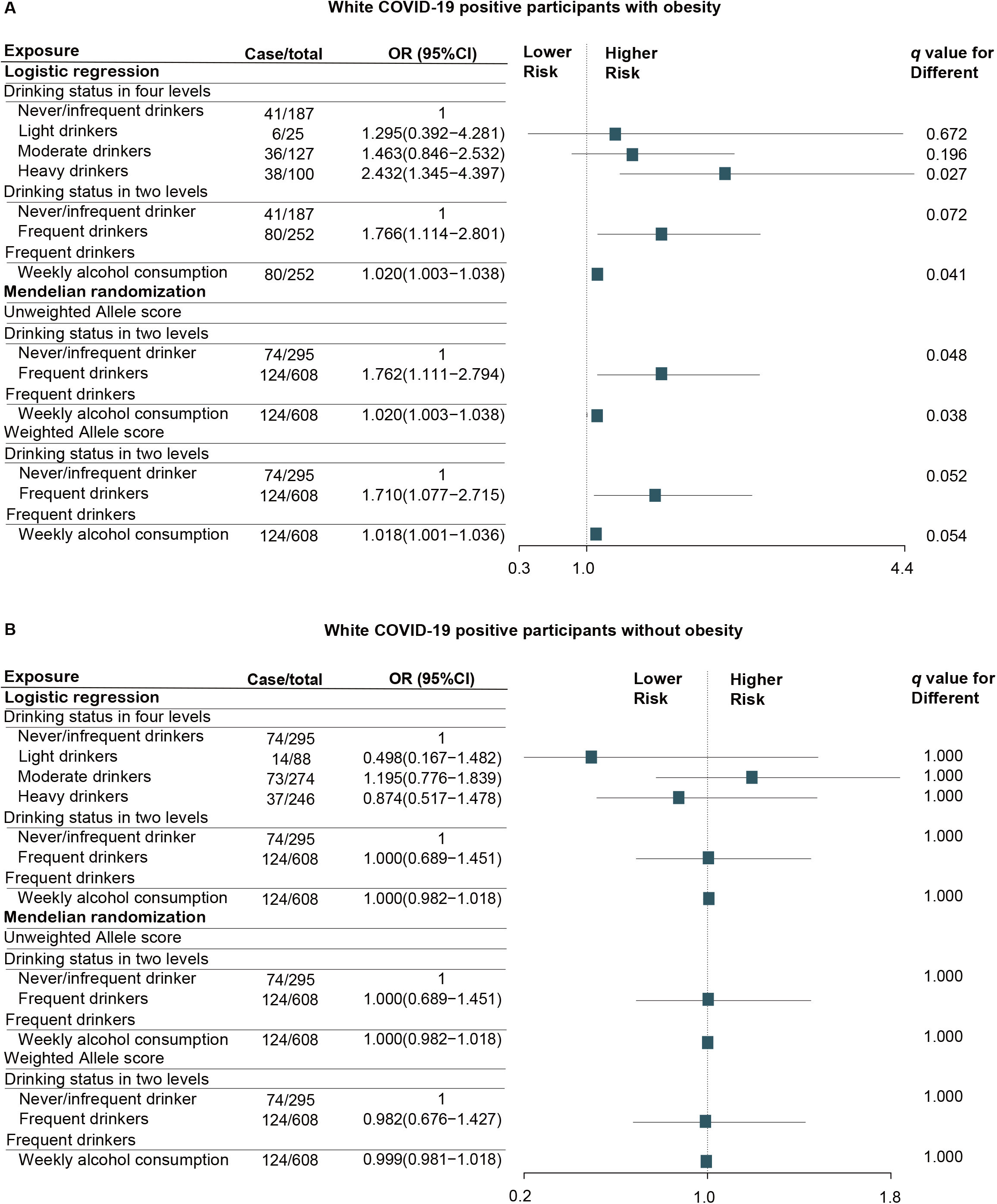
Logistic regression and Mendelian randomization analyses of the association of alcohol consumption with the risk of severe clinical outcomes in white COVID-19 positive patients with (A) and without obesity (B). Analyses were performed in PSM cohort. Matching factors for PSM including age, sex, BMI categories, current smoking status, alcohol related diseases, asthma, emphysema, COPD, bronchitis/bronchiectasis, esophagitis, gastritis/duodenitis, peptic ulcer, GERD, hypertensive, chronic ischemic heart disease, heart failure, diabetes, dementia, renal failure, liver cirrhosis and/or liver failure, tumor and AIDS. q-value was calculated by false discovery rate (FDR) method. Abbreviation: OR, odds ratio; HR, hazard ratio; CI, confidence interval; PSM, propensity score matching; BMI, body mass index; GERD, gastroesophageal reflux disease; COPD, chronic obstructive pulmonary disease; AIDS, acquired immunodeficiency syndrome.

## Discussion

Using the UK Biobank cohort, we investigated whether alcohol consumption increased susceptibility to SARS-CoV-2 infection among 12,937 white participants who have been tested for SARS-COV-2, as well as whether there were worse outcomes among 1,570 patients with COVID-19 using regression analyses and Mendelian randomization analysis. We found that alcohol consumption did not increase susceptibility to SARS-CoV-2 infection; however, frequent drinking, especially heavy drinking, was associated with worse outcomes of COVID-19 in patients with obesity, but not non-obese patients. Notably, the risk of worse clinical outcomes in frequent drinkers with obesity increased slightly with the average amount of alcohol consumed weekly.

### Possible explanations for an interaction between alcohol consumption and worse outcomes of COVID-19 in patients with obesity

According to our findings, frequent drinking was associated with poor outcomes of COVID-19 in patients with obesity, but not non-obese, patients, suggesting the potential interactions between alcohol consumption and obesity to cause rapid progression of COVID-19. Consistent with a previous report[34], our study found that patients with obesity were more susceptible to SARS-CoV-2 infection (OR=1.260, 95%CI 1.092-1.453; *P*=.002) and had a higher risk of death (HR=1.561, 95%CI=1.122-2.171; *P*=.008) from COVID-19. The high expression of angiotensin-converting enzyme 2 receptor (ACE2) in adipocytes of patients with obesity may promote the entry of SARS CoV2 into host cells and turn adipose tissue into a potential target and reservoir of SARS CoV2[35]. Moreover, adipose tissue, a major source of pro-inflammatory chemokines, cytokines, and adipokines, plays an important role in mediating inflammatory responses. Patients with obesity have higher concentrations of circulating tumor necrosis factor-alpha (TNF-a), interleukin-6, and C-reactive protein (CRP); this low grade chronic inflammatory state may be involved in initiating cytokine storms in patients with COVID-19[34].

Alcohol consumption is associated with an increased risk of ARDS[12], which is a common manifestation of severe COVID-19. This is likely due to the dysregulation of both immune and non-immune host defense mechanisms in the airways, resulting in alveolar epithelial barrier dysfunction and alveolar macrophage immune dysregulation in response to heavy alcohol consumption[9, 12]. However, the mechanisms for the potential interaction between alcohol and obesity in the progression of COVID-19 are not well understood. Previous work has found that chronic ethanol exposure in murine models impacts adipose tissue, phenocopying obesity in many aspects. Chronic ethanol feeding decreases glucose uptake by adipose tissue, increases immune cell infiltration and expression of inflammatory cytokines and adipokines[36]. Data from pre-clinical models also demonstrates that binge drinking and obesity synergistically induces steatohepatitis and fibrosis in the liver of mice via the induction of hepatic chemokines, inflammatory cytokines and neutrophil infiltration[37, 38]. Taken together, alcohol intake and obesity may affect the progression of COVID-19 via impact on adipose tissue and inflammatory responses; the additive and/or synergistic effects of obesity and alcohol may lead to a significant deterioration of COVID-19 patients. However, mechanisms for the potential interactions between alcohol and obesity in the progression of COVID-19 are not well understood.

### Strengths and Limitations

One of main advantages of this study is the application of Mendelian randomization to evaluate the causal relationships between alcohol consumption and the risk of SARS CoV2 infection and the severity of COVID-19 outcomes. To our knowledge, the associations between alcohol and risk of COVID-19 have not been previously explored using Mendelian randomization. We generated weighted and unweighted allele scores using three SNPs rs1229984 (ADH1B), rs1260326 (GCKR), and rs13107325 (SLC39A8) and applied the weighted and unweighted allele scores as the instrumental variables to assess the effect of alcohol consumption on outcomes. These three SNPs were randomly distributed among the white participants and were not associated with other confounders that may affect outcomes. However, rs1229984, rs1260326, and rs13107325 were significantly correlated with obesity. This issue was well addressed by our separate analysis based on whether the white participants were obese or not. In addition, the Mendelian randomization approach more robustly handles measurement error and reverse causality compared to traditional observational approaches. Another major advantage of our study is the detailed data in a larger population-based prospective cohort including genotype data, alcohol consumption, and potential confounding risk factors. Almost all the confounding factors that have been reported to be associated with the risk of COVID-19 are available in the UK Biobank. Therefore, we were able to balance these factors in non-drinkers and frequent drinkers using PSM to better assess the specific effect of alcohol consumption on the risk of COVID-19 in the traditional observational study and Mendelian randomization study.

This study also has several limitations. First, since the majority of participants in the study were British and Irish descents (93%) and only 195 non-white participants were infected with SARS CoV2, we could not investigate the impact of alcohol consumption on the risk of worse outcomes in non-white participants with COVID-19 using Mendelian randomization analysis. However, according to the results of the Chi-square test, we found heavy drinkers had a higher mortality than non-heavy drinkers (57.1% vs. 18.0%, *P*=.038) in the non-white group with obesity, which was not observed in non-obese group (20.0% vs. 8.0%, *P*=0.220). Second, we are unable to assess the effect of changes in the alcohol consumption since subjects (n=501,608) were enrolled in the UK Biobank in 2006-2010. Although the self-reported alcohol consumption data of some participants were updated in 2012 (n=20,336), 2014 (n=48,340) and 2019 (n=3,081), the number of these patients with COVID-19 was relatively small and the corresponding statistical analysis could not be performed. Only 5 COVID-19 positive patients had their alcohol consumption data updated in 2019 and there was no significant change in their drinking patterns (data not shown). Finally, due to the limitations of the data in the UK Biobank, we could not assess the impact of alcohol consumption on the symptoms and some severe clinical outcomes (requirement of oxygen therapy, glucocorticoid therapy, and administration of invasive ventilation) of patients. To minimize this limitation, we classified ICU admission and death as the severe clinical outcomes of COVID-19 positive patients and found heavy drinkers had a higher risk of admission to ICU and death in patients with obesity compared to non-drinkers.

## Conclusions

Despite the limitations, our study was the first to find that alcohol consumption, especially heavy drinking, is associated with a higher risk of suffering worse COVID-19 clinical outcomes in patients with obesity through both traditional regression analyses and Mendelian randomization analyses. In addition, alcohol consumption was not associated with either increased or decreased risk of SARS CoV2 infection. Our findings could help people understand the relationship between alcohol consumption and COVID-19, especially those who may drink excessively in the mistaken belief that alcohol consumption reduces the risk of SARS CoV2 infection[18, 19]. Moreover, due to the possible interactions between alcohol consumption and obesity in the progression of COVID-19, physicians may need to adjust and develop appropriate management and treatment strategies for COVID-19 positive patients who consume alcohol and are obese.

## Data Availability

This study used data from the UK Biobank (application number 59473). For details please contact. All other data are contained in the article and its supplementary information or available upon reasonable request.

https://www.ukbiobank.ac.uk/

## Acknowledgments

This study has been conducted using the UK Biobank Resource (application number 59473). The views expressed are those of the authors and not necessarily those of the National Institute for Health Research (NIHR) or the Department of Health and Social Care.

## Data availability

### Abbreviation

COVID-19: coronavirus disease 2019
SARS-CoV-2: severe acute respiratory syndrome coronavirus-2
HR: hazard ratio
OR: odds ratio
CI: confidence interval
N: number of participants
ADH1B: alcohol dehydrogenase 1B
BMI: body mass index
GERD: gastroesophageal reflux disease
COPD: chronic obstructive pulmonary disease
AIDS: acquired immunodeficiency syndrome
ICU: intensive care unit
ARDS: acute respiratory distress syndrome
RSV: respiratory syncytial virus
FDR: false discovery rate
SNP: single nucleotide polymorphism
PSM: propensity score matching
RAF: risk allele frequency
GERA: Genetic Epidemiology Research in Adult Health and Aging
ADH1B: alcohol dehydrogenase 1B
GCKR: glucokinase regulator
SLC39A8: solute carrier family 39 member 8

## Supporting information

**Table S1**. Detailed information about the genetic variations of ADH1B/ SLC39A8/ GCKR included in this study.

**Table S2**. Diseases diagnosis codes used by the UK Biobank.

**Table S3**. Association of observed confounders with alcohol consumption.

**Table S4**. Characteristics of participants by rs1229984, rs1260326, and rs13107325 genotypes.

**Table S5**. Association of genetic variations of ADH1B/SLC39A8/GCKR with outcomes of interest in white participants.

**Table S6**. Logistic/Cox regression and Mendelian randomization analyses of the associations of alcohol consumption with the risk of SARS-CoV-2 infection and the risk of death of COVID-19 in white participants who were overweight but not obese.

**Figure S1**. Association of combined ADH1B, SLC39A8, GCKR fast-allele score with average alcohol consumption levels (standard drink/weekly) in whole participants (A) and percentages of heavy-drinkers (B) in whole participants by unweighted allele score. The amount of alcohol consumed by non-drinkers was defined as zero.

