## Supplemental Table 1-6 for "Alcohol Consumption is Associated with Poor Prognosis in Obese Patients with COVID-19: a Mendelian Randomization Study using UK Biobank"

**Table S1.** **Detailed information about the genetic variations of ADH1B/ SLC39A8/GCKR included in this study ^1^.**

| **SNP** | **Locus** | **Risk allele** | **Other allele** | **UK Biobank heavy alcohol drinker status (cases versus controls)** | | | **GERA database drinks/ week (among drinkers)** | |
| --- | --- | --- | --- | --- | --- | --- | --- | --- |
|  |  |  |  | **RAF** | **OR (95%CI)** | **P value** | **β(SE)** | **P value** |
| **rs1229984** | ADH1B | C | T | 0.98 | 1.58(1.48-1.70) | 3.30×10^−36^ | 0.187(0.016) | 2.9×10^−32^ |
| **rs1260326** | GCKR | C | T | 0.612 | 1.06(1.04-1.08) | 2.60×10^−8^ | 0.033(0.029) | 1.1×10^−6^ |
| **rs13107325** | SLC39A8 | C | T | 0.928 | 1.12(1.08-1.16) | 1.60× 10^−8^ | 0.029(0.013) | 0.0249 |

**Abbreviation:** OR, odds ratio; CI, confidence interval; RAF, risk allele frequency; GERA, Genetic Epidemiology Research in Adult Health and Aging; ADH1B, alcohol dehydrogenase 1B; GCKR, glucokinase regulator; SLC39A8, solute carrier family 39 member 8.

| **Table S2. Disease diagnosis codes used by the UK Biobank** | |
| --- | --- |
| **Diseases** | **Diagnosis code** |
| **Alcohol related diseases** |  |
| Alcohol use disorder | F10.0-F10.3 |
| Alcohol liver diseases | K70.0-K70.9 |
| Alcohol pancreatitis | K85.2 and K86.0 |
| Alcoholic gastritis | K29.2 |
| Alcoholic cardiomyopathy | I42.6 |
| Alcoholic psychosis | F10.3-F10.9 |
| Alcoholic myopathy | G72.1 |
| Alcoholic polyneuropathy, | G62.1 |
| Degeneration of the nervous system due to alcohol | G31.2 |
| **Upper gastrointestinal diseases** |  |
| Oesophagitis | K22.0-K22.9 |
| GERD | K21.0, K21.9 |
| Peptic ulcer | K25.0-K28.9 |
| Gastritis/duodenitis | K29.0-K29.8 |
| **Chronic lower respiratory diseases** |  |
| COPD | J44.0, J44.1, J44.8, J44.9 |
| Emphysema | J43.0-J43.2, J43.8, J43.9 |
| Bronchitis/Bronchiectasis | J40, J41.0, J41.1, J42, J47 |
| Asthma | J45.0, J45.1, J45.8, J45.9, J46 |
| **Chronic heart diseases** |  |
| Heart failure | I50.0-I50.9 |
| Hypertensive | I10-I15.9 |
| Chronic ischaemic heart disease | I23.0-I25.9 |
| **Diabetes mellitus** | E10.0-E14.9 |
| **Dementia** | F00.0-F03 |
| **Liver cirrhosis and/or liver failure** | R18, K70.3, K71.7, K72.9, K74.3-K74.6, K76.6, K76.7, K70.4, K72, K72.0, K72.1, K72.9 |
| **Renal failure** | N17.0-N19 |
| **AIDS** | B20.0-B24 |

**Abbreviation:** GERD, gastroesophageal reflux disease; COPD, chronic obstructive pulmonary disease; AIDS, acquired immunodeficiency syndrome.

**Table S3. Association of observed confounders with alcohol consumption**

| **Variables** | **Non-drinkers (n=4496)** | **Frequent drinkers (n=8441)** | **Light drinkers (n=1156)** | **Moderate drinkers (n=3795)** | **Heavy drinkers (n=3490)** | **q-value** |
| --- | --- | --- | --- | --- | --- | --- |
| **Age (years), n(%)** |  |  |  |  |  | 0.002 |
| <65 | 1379(30.7) | 2362(28.0) | 330(28.5) | 1012(26.7) | 1020(29.2) |  |
| ≥65 | 3117(69.3) | 6079(72.0) | 826(71.5) | 2783(73.3) | 2470(70.8) |  |
| **Male, n(%)** | 1648(36.7) | 4687(55.5) | 414(35.8) | 2502(65.9) | 1771(50.7) | <0.001 |
| **Race, n(%)** |  |  |  |  |  | <0.001 |
| No white | 622(13.9) | 286(3.4) | 56(4.9) | 149(3.9) | 81(2.3) |  |
| White | 3851(86.1) | 8131(96.6) | 1090(95.1) | 3636(96.1) | 3405(97.7) |  |
| **BMI categories, n(%)** |  |  |  |  |  | <0.001 |
| Normal weight (18.5-24.9) | 1059(24.2) | 2363(28.6) | 365(32.2) | 1008(27.2) | 990(29.0) |  |
| Underweight (<18.5) | 26(0.6) | 38(0.5) | 12(1.1) | 10(0.3) | 16(0.5) |  |
| Overweight (25-29.9) | 1616(37.0) | 3616(43.8) | 459(40.5) | 1691(45.6) | 1466(43.0) |  |
| Obesity (≥30) | 1671(38.2) | 2232(27.1) | 296(26.1) | 999(26.9) | 937(27.5) |  |
| **Blood type, n(%)** |  |  |  |  |  | 0.002 |
| OO | 1853(41.3) | 3605(42.8) | 525(45.6) | 1610(42.5） | 1470(42.1) | 0.002 |
| AA+AO | 181(4.0) | 266(3.2) | 45(3.9) | 123(3.2) | 98(2.8) |  |
| BB+BO | 514(11.5） | 815(9.7) | 108(9.4) | 371(9.8) | 336(9.6) |  |
| AB | 1941(43.2) | 3745(44.4) | 473(41.1) | 1688(44.5) | 1584(45.4) |  |
| **Current smoking, n(%)** |  |  |  |  |  | <0.001 |
| No | 3908(87.0) | 7421(88.0) | 1064(92.1) | 3445(90.8) | 2912(83.5) |  |
| Only occasionally | 122(2.7) | 274(3.2) | 23(2.0) | 104(2.7) | 147(4.2) |  |
| Most or all days | 461(10.3) | 741(8.8) | 68(5.9) | 244(6.4) | 429(12.3) |  |
| **Comorbidities, n(%)** |  |  |  |  |  |  |
| **Upper gastrointestinal diseases** |  |  |  |  |  |  |
| Oesophagitis | 239(5.3) | 361(4.3) | 36(3.1) | 175(4.6) | 150(4.3) | 0.011 |
| GERD | 601(13.4) | 836(9.9) | 121(10.5) | 356(9.4) | 359(10.3) | <0.001 |
| Peptic ulcer | 187(4.2) | 266(3.2) | 28(2.4) | 117(3.1) | 121(3.5) | 0.011 |
| Gastritis/duodenitis | 664(14.8) | 868(10.3) | 126(10.9) | 374(9.9) | 369(10.5) | <0.001 |
| **Chronic lower respiratory diseases** |  |  |  |  |  |  |
| COPD | 340(7.6) | 446(5.3) | 43(3.7) | 193(5.1) | 210(6.0) | <0.001 |
| Emphysema | 63(1.4) | 84(1.0) | 8(0.7) | 44(1.2) | 32(0.9) | 0.114 |
| Bronchitis/Bronchiectasis | 78(1.7) | 126(1.5) | 25(2.2) | 50(1.3) | 51(1.5） | 0.177 |
| Asthma | 640(14.2) | 821(9.7) | 115(9.9) | 346(9.1) | 360(10.3) | <0.001 |
| **Chronic heart diseases** |  |  |  |  |  |  |
| Heart failure | 216(4.8) | 290(3.4) | 29(2.5) | 139(3.7) | 122(3.5) | 0.001 |
| Hypertensive | 1839(40.9） | 2857(33.8) | 372(32.2) | 1291(34.0) | 1194(34.2） | <0.001 |
| Chronic ischaemic heart disease | 699(15.5) | 1038(12.3) | 121(10.5) | 514(13.5) | 403(11.5) | <0.001 |
| **Diabetes mellitus** | 796(17.7) | 770(9.1) | 117(10.1) | 393(10.4) | 260(7.4) | <0.001 |
| **Liver cirrhosis and/or liver failure** | 43(1.0) | 48(0.6) | 6(0.5) | 15(0.4) | 27(0.8) | 0.023 |
| **Renal failure** | 408(9.1) | 541(6.4) | 78(6.7) | 257(6.8) | 206(5.9) | <0.001 |
| **Insomnia** | 3578(79.7) | 6486(76.9) | 885(77.0) | 2831(74.6) | 2770(79.4) | <0.001 |
| **Dementia** | 37(0.8） | 56(0.7) | 13(1.1) | 22(0.6) | 21(0.6) | 0.183 |
| **Tumor** | 461(10.3) | 862(10.2) | 114(9.9) | 391(10.3) | 357(10.2) | 1.000 |
| **AIDS** | 5(0.1) | 8(0.1) | 2(0.2) | 5(0.1) | 1(0.03) | 1.000 |
| **COVID-19 positivity, n(%)** | 643(14.3) | 927(11.0) | 126(10.9) | 433(11.4) | 368(10.5) | <0.001 |
| **COVID-19 positive patients, n(%)** |  |  |  |  |  |  |
| **Death** | 115(17.9) | 172(18.6) | 14(11.1) | 90(20.8) | 68(18.5) | 0.430 |
| **ICU admission and death** | 147(22.9) | 219(23.6) | 21(16.7) | 115(26.6) | 83(22.6) | 0.581 |

**q-value** was calculated by false discovery rate (FDR) method.

**Abbreviation:** BMI, body mass index; GERD, gastroesophageal reflux disease; COPD, chronic obstructive pulmonary disease; AIDS, acquired immunodeficiency syndrome; COVID-19, coronavirus disease 2019; ICU, intensive care unit.

**Table S4. Characteristics of participants by rs1229984, rs1260326, and rs13107325 genotypes.**

|  | **ADH1B rs1229984** | | | **SLC39A8 rs13107325** | | | **GCKR rs1260326** | | |
| --- | --- | --- | --- | --- | --- | --- | --- | --- | --- |
| **Variables** | **1/1 (slow) n=12172** | **1/2 or 2/2 (fast) n=765** | **q-value** | **1/1 (slow) n=11160** | **1/2 or 2/2 (fast) n=1756** | **q-value** | **1/1 (slow) n=4947** | **1/2 or 2/2 (fast) n=7920** | **q-value** |
| **Age (years), n(%)** |  |  | 0.883 |  |  | 0.788 |  |  | 1.000 |
| <65 | 3500(28.8) | 241(31.5) |  | 3274(29.3) | 463(26.4) |  | 1461(29.5) | 2252(28.4) |  |
| ≥65 | 8672(71.2) | 524(68.5) |  | 7886(70.7) | 1293(73.6) |  | 3486(70.5) | 5668(71.6) |  |
| **Male, n(%)** | 5977(49.1) | 358(46.8) | 0.808 | 5443(48.4) | 881(50.2) | 0.803 | 2452(49.6) | 3856(48.7) | 0.224 |
| **Race, n(%)** |  |  | <0.001 |  |  | <0.001 |  |  | <0.001 |
| No white | 767(6.3) | 141(18.5) |  | 879(7.9) | 28(1.6) |  | 512(10.4) | 380(4.8) |  |
| White | 11359(93.7) | 623(81.5) |  | 10252(92.1) | 1720(98.4) |  | 4416(89.6) | 7514(95.2) |  |
| **BMI categories, n(%)** |  |  |  |  |  |  |  |  |  |
| Normal weight (18.5-24.9) | 3201(27.0) | 221(29.4) |  | 3019(27.7) | 396(23.0) |  | 1263(26.3) | 2137(27.6) |  |
| Underweight (<18.5) | 59(0.5) | 5(0.7) | 1.000 | 54(0.5) | 10(0.6) | 0.756 | 23(0.5) | 40(0.5) | 0.989 |
| Overweight (25-29.9) | 4891(41.2) | 341(45.4) | 1.000 | 4460(41.0) | 762(44.3) | 0.002 | 1946(40.5) | 3267(42.2) | 0.949 |
| Obesity (≥30) | 3719(31.3) | 184(24.5) | 0.058 | 3350(30.8) | 551(32.1) | 0.007 | 1577(32.8) | 2299(29.7) | 0.068 |
| **Blood type, n(%)** |  |  |  |  |  |  |  |  |  |
| OO | 5128(42.2) | 330(43.1) |  | 4706(42.2) | 742(42.4) |  | 2098(42.5) | 3331(42.1) |  |
| AA+AO | 416(3.4) | 31(4.1) | 0.913 | 388(3.5) | 58(3.3) | 0.972 | 169(3.4) | 274(3.5) | 0.905 |
| BB+BO | 1221(10.0) | 108(14.1) | 0.770 | 1155(10.4) | 172(9.8) | 0.853 | 520(10.5) | 800(10.1) | 0.530 |
| AB | 5390(44.3) | 296(38.7) | 0.925 | 4900(44.0) | 780(44.5) | 0.903 | 2154(43.6) | 3504(44.3) | 1.000 |
| **Current smoking, n(%)** |  |  |  |  |  |  |  |  |  |
| No | 10656(87.6) | 673(88.0) |  | 9775(87.6) | 1536(87.6) |  | 4318(87.3) | 6947(87.8) |  |
| Only occasionally | 369(3.0) | 27(3.5) | 1.000 | 351(3.1) | 44(2.5) | 0.788 | 157(3.2) | 236(3.0) | 0.972 |
| Most or all days | 1137(9.3) | 65(8.5) | 0.873 | 1027(9.2) | 173(9.9) | 0.812 | 470(9.5) | 729(9.2) | 0.501 |
| **Comorbidities, n(%)** |  |  |  |  |  |  |  |  |  |
| **Upper gastrointestinal diseases** | |  |  |  |  |  |  |  |  |
| Oesophagitis | 579(4.8) | 21(2.7) | 0.812 | 518(4.6) | 81(4.6) | 0.864 | 188(3.8) | 409(5.2) | 0.015 |
| GERD | 1363(11.2) | 74(9.7) | 0.997 | 1226(11.0) | 207(11.8) | 0.854 | 559(11.3) | 871(11.0) | 0.561 |
| Peptic ulcer | 428(3.5) | 25(3.3) | 1.000 | 396(3.5) | 57(3.2) | 0.564 | 175(3.5) | 277(3.5) | 1.000 |
| Gastritis/duodenitis | 1458(12.0) | 74(9.7) | 0.812 | 1311(11.7) | 218(12.4) | 0.584 | 570(11.5) | 953(12.0) | 0.991 |
| **Chronic lower respiratory diseases** | |  |  |  |  |  |  |  |  |
| COPD | 753(6.2) | 33(4.3) | 0.843 | 677(6.1) | 109(6.2) | 0.597 | 295(6.0) | 489(6.2) | 1.000 |
| Emphysema | 141(1.2) | 6(0.8) | 0.971 | 118(1.1) | 28(1.6) | 0.412 | 50(1.0) | 97(1.2) | 0.493 |
| Bronchitis/Bronchiectasis | 196(1,6) | 8(1.0) | 0.919 | 165(1.5) | 39(2.2) | 0.232 | 71(1.4) | 131(1.7) | 1.000 |
| Asthma | 1383(11.4) | 78(10.2) | 0.915 | 1259(11.3) | 199(11.3) | 0.972 | 538(10.9) | 914(11.5) | 0.464 |
| **Chronic heart diseases** |  |  |  |  |  |  |  |  |  |
| Heart failure | 471(3.9) | 35(4.6) | 0.667 | 440(3.9) | 64(3.6) | 0.812 | 184(3.7) | 318(4.0) | 0.510 |
| Hypertensive | 4444(36.5) | 252(32.9) | 0.750 | 4065(36.4) | 625(35.6) | 0.527 | 1815(36.7) | 2857(36.1) | 1.000 |
| Chronic ischaemic heart disease | 1640(13.5) | 97(12.7) | 0.942 | 1496(13.4) | 239(13.6) | 0.930 | 628(12.7) | 1101(13.9) | 0.087 |
| **Diabetes mellitus** | 1475(12.1) | 91(11.9) | 0.936 | 1349(12.1) | 215(12.2) | 0.859 | 662(13.4) | 893(11.3) | 0.151 |
| **Serious liver diseases** | 88(0.7) | 3(0.4) | 0.887 | 79(0.7) | 12(0.7) | 0.902 | 29(0.6) | 62(0.8) | 0.474 |
| **Renal failure** | 897(7.4) | 52(6.8) | 0.926 | 807(7.2) | 139(7.9) | 0.722 | 388(7.8) | 556(7.0) | 0.515 |
| **Insomnia** | 9463(77.8) | 601(78.6) | 0.822 | 8675(77.8) | 1373(78.2) | 0.894 | 3887(78.7) | 6128(77.4) | 0.203 |
| **Dementia** | 91(0.7) | 2(0.3) | 0.875 | 80(0.7) | 13(0.7) | 0.931 | 44(0.9) | 49(0.6) | 0.164 |
| **Tumor** | 1246(10.2) | 77(10.1) | 0.897 | 1143(10.2) | 177(10.1) | 0.815 | 505(10.2) | 815(10.3) | 0.635 |
| **AIDS** | 11(0.1) | 2(0.3) | 0.719 | 12(0.1) | 1(0.1) | 0.827 | 4(0.1) | 8(0.1) | 1.000 |

**q-value** was calculated by false discovery rate (FDR) method.

**Abbreviation:** BMI, body mass index; GERD, gastroesophageal reflux disease; COPD, chronic obstructive pulmonary disease; AIDS, acquired immunodeficiency syndrome.

**Table S5. Association of genetic variations of ADH1B/SLC39A8/GCKR with outcomes of interest in white participants**

| **Instrumental variables** | **OR of SARS-CoV-2 infection** | **HR of death in COVID-19 positive patients** | **OR of ICU admission and death in COVID-19 positive patients** |
| --- | --- | --- | --- |
| **ADH1B one or two fast alleles vs. none** | 0.965(0.746-1.247) | 0.715(0.367-1.390) | 0.592(0.307-1.143) |
| **P-value** | 0.783 | 0.322 | 0.118 |
| **SLC39A8 one or two fast alleles vs. none** | 0.984(0.837-1.156) | 1.116(0.797-1.565) | 1.163(0.822-1.646) |
| **P-value** | 0.843 | 0.522 | 0.393 |
| **GCKR one or two fast alleles vs. none** | 0.994(0.884-1.117) | 0.899(0.699-1.156) | 0.839(0.650-1.082) |
| **P-value** | 0.918 | 0.407 | 0.176 |
| **Unweighted allele score** | 0.996(0.930-1.068) | 0.967(0.832-1.124) | 0.917(0.787-1.069) |
| **P-value** | 0.919 | 0.661 | 0.269 |
| **Weighted allele score** | 0.959(0.310-2.965) | 0.318(0.023-4.479) | 0.113(0.008-1.633) |
| **P-value** | 0.942 | 0.396 | 0.110 |

**Abbreviation:** OR, odds ratio; HR, hazard ratio; CI, confidence interval; ICU, intensive care unit.

**Table S6. Logistic/Cox regression and Mendelian randomization analyses of the associations of alcohol consumption with the risk of SARS-CoV-2 infection and the risk of death of COVID-19 in white participants who were overweight but not obese.**

| **Exposure and outcomes** | **The risk of SARS-CoV-2 infection (Overweight)** | | | **The risk of death (Overweight)** | | |
| --- | --- | --- | --- | --- | --- | --- |
|  | **Case/total** | **OR (95%CI)** | **q-value** | **Case/total** | **HR (95%CI)** | **q-value** |
| **Logistic and Cox regression** |  |  |  |  |  |  |
| **Drinking status in four levels** |  |  |  |  |  |  |
| Never/infrequent drinkers | 181/1383 | 1 |  | 34/181 | 1 |  |
| Light drinkers | 52/431 | 0.881(0.484-1.604) | 0.763 | 8/52 | 0.347(0.047-2.538) | 1.000 |
| Moderate drinkers | 193/1622 | 0.967(0.748-1.250) | 0.797 | 43/193 | 1.204(0.720-2.013) | 1.000 |
| Heavy drinkers | 142/1433 | 0.766(0.562-1.044) | 0.828 | 18/142 | 0.963(0.507-1.831) | 0.909 |
| **Drinking status in two levels** |  |  |  |  |  |  |
| Never/infrequent drinker | 181/1383 | 1 | 0.486 | 34/181 | 1 | 1.000 |
| Frequent drinkers | 387/3486 | 0.883(0.708-1.101) |  | 69/387 | 1.054(0.664-1.671) |  |
| **Frequent drinkers** |  |  |  |  |  |  |
| Weekly alcohol consumption | 387/3486 | 0.907(0.740-1.113) | 0.525 | 69/387 | 1.189(0.789-1.772) | 1.000 |
| **Mendelian randomization** |  |  |  |  |  |  |
| **Unweighted allele score** |  |  |  |  |  |  |
| **Drinking status in two levels** |  |  |  |  |  |  |
| Never/infrequent drinker | 181/1383 | 1 | 0.601 | 34/181 | 1 | 1.000 |
| Frequent drinkers | 387/3486 | 0.882(0.707-1.101) |  | 69/387 | 1.053(0.664-1.669) |  |
| **Frequent drinkers** |  |  |  |  |  |  |
| Weekly alcohol consumption | 387/3486 | 0.992(0.981-1.003) | 0.621 | 69/387 | 1.005(0.96-1.025) | 1.000 |
| **Weighted allele score** |  |  |  |  |  |  |
| **Drinking status in two levels** |  |  |  |  |  |  |
| Never/infrequent drinker | 181/1383 | 1 | 0.469 | 34/181 | 1 | 0.978 |
| Frequent drinkers | 387/3486 | 0.902(0.723-1.127) |  | 69/387 | 1.040(0.655-1.649) |  |
| **Frequent drinkers** |  |  |  |  |  |  |
| Weekly alcohol consumption | 387/3486 | 0.993(0.982-1.004) | 0.567 | 69/387 | 1.005(0.985-1.024) | 1.000 |

**Analyses** were performed in PSM cohort. Matching factors for PSM including age, sex, BMI categories, current smoking status, alcohol related diseases, asthma, emphysema, COPD, bronchitis/bronchiectasis, esophagitis, gastritis/duodenitis, peptic ulcer, GERD, hypertensive, chronic ischemic heart disease, heart failure, diabetes, dementia, renal failure, liver cirrhosis and/or liver failure, tumor and AIDS.

**q-value** was calculated by false discovery rate (FDR) method.

**Abbreviation:** OR, odds ratio; HR, hazard ratio; CI, confidence interval; PSM, propensity score matching; BMI, body mass index; GERD, gastroesophageal reflux disease; COPD, chronic obstructive pulmonary disease; AIDS, acquired immunodeficiency syndrome.

**Figure S1**

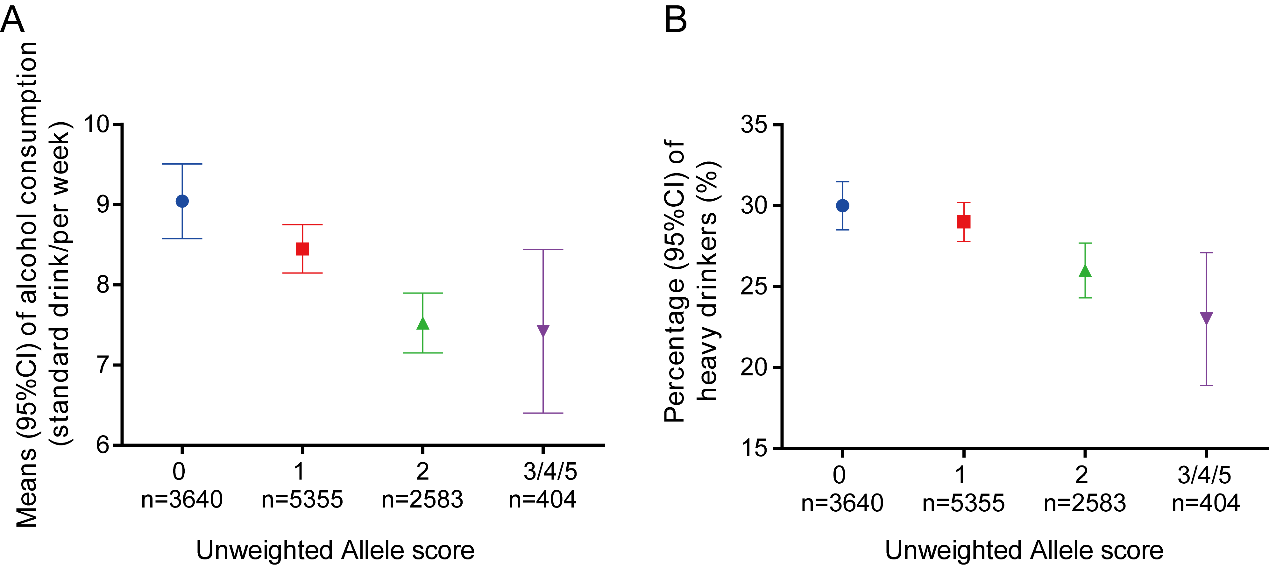

Figure S1. Association of combined ADH1B, SLC39A8, GCKR fast-allele score with average alcohol consumption levels (standard drink/weekly) in whole participants (A) and percentages of heavy-drinkers (B) in whole participants by unweighted allele score. The amount of alcohol consumed by non-drinkers was defined as zero.
